# Mortality among Care Home Residents in England during the first and second waves of the COVID-19 pandemic: an analysis of 4.3 million adults over the age of 65

**DOI:** 10.1101/2021.07.07.21253295

**Authors:** Anna Schultze, Emily Nightingale, David Evans, William Hulme, Alicia Rosello, Chris Bates, Jonathan Cockburn, Brian MacKenna, Helen J Curtis, Caroline E Morton, Richard Croker, Seb Bacon, Helen I McDonald, Christopher T Rentsch, Krishnan Bhaskaran, Rohini Mathur, Laurie A Tomlinson, Elizabeth J Williamson, Harriet Forbes, John Tazare, Daniel Grint, Alex J Walker, Peter Inglesby, Nicholas J DeVito, Amir Mehrkar, George Hickman, Simon Davy, Tom Ward, Louis Fisher, Amelia CA Green, Kevin Wing, Angel YS Wong, Robert McManus, John Parry, Frank Hester, Sam Harper, Stephen JW Evans, Ian J Douglas, Liam Smeeth, Rosalind M Eggo, Ben Goldacre, David A Leon

**Author notes:** Joint Contribution (Ben Goldacre, David A Leon). Corresponding Dr Anna Schultze, Department of Non-Communicable Disease Epidemiology, London School of Hygiene and Tropical Medicine, Keppel Street, London WC1E 7HT.

## Abstract

**Background:** Residents in care homes have been severely impacted by the COVID-19 pandemic. We describe trends in risk of mortality among care home residents compared to residents in private homes in England.

**Methods:** On behalf of NHS England, we used OpenSAFELY-TPP, an analytics platform running across the linked electronic health records of approximately a third of the English population, to calculate monthly age-standardised risks of death due to all causes and COVID-19 among adults aged >=65 years between 1/2/2019 and 31/03/2021. Care home residents were identified using linkage to the Care and Quality Commission.

**Findings:** We included 4,329,078 people aged 65 years or older on the 1st of February 2019, 2.2% of whom were classified as residing in a care or nursing home. Age-standardised mortality risks were approximately 10 times higher among care home residents compared to non-residents in February 2019 residents (CMF = 10.59, 95%CI = 9.51, 11.81 among women, CMF = 10.82, 95%CI = 9.89, 11.84 among men). This increased to more than 17 times in April 2020 (CMF = 17.52, 95%CI = 16.38, 18.74 among women, CMF = 18.12, 95%CI = 17.17 – 19.12 among men) before returning to pre-pandemic levels in June 2020. CMFs did not increase during the second wave, despite a rise in the absolute age-standardised COVID-19 mortality risks.

**Interpretation:** The first COVID-19 wave had a disproportionate impact on care home residents in England compared to older private home residents. A degree of immunity, improved protective measures or changes in the underlying frailty of the populations may explain the lack of an increase in the relative mortality risks during the second wave. The care home population should be prioritised for measures aimed at controlling the spread of COVID-19.

**Funding:** Medical Research Council MR/V015737/1

## Background

The COVID-19 pandemic has had a major adverse effect on the residents of care homes, their relatives, and carers in the UK and in many other countries (1). At the end of the first wave in England and Wales, the Office for National Statistics (ONS) estimated that almost a third of all deaths occurring among care home residents were due to COVID-19 (2). The cumulative number of COVID-19 deaths among care home residents at that point, just over 19,000, represented approximately 40% of all COVID-19 deaths in England and Wales (3). However, this is likely an underestimate given the low levels of testing in care homes at the time. The Health Foundation estimates that there were approximately 10,000 additional so-called “excess” deaths among care home residents in England alone during the first wave (3). Later research has shown that the vast majority of excess care home deaths in England and Scotland have occurred in care homes where there were COVID-19 outbreaks, indicating that these deaths were likely due to undiagnosed COVID-19 (4,5).

The impact of the COVID-19 pandemic on the risk of death among care home residents in England has not yet been comprehensively investigated. This is in part because of the absence of any national registry of care home residents. Working on behalf of NHS England, we used the OpenSAFELY data analytics platform to investigate the risk of death of individuals in care homes compared to that of individuals in private residences from pre-pandemic to early 2021. Describing and understanding these differences and trends is an essential component of learning the lessons of COVID-19 so we are better prepared in the future.

## Methods

### Data Source

Primary care records managed by the GP software provider TPP were linked to ONS death data through OpenSAFELY, a data analytics platform created by our team on behalf of NHS England to address urgent COVID-19 research questions (https://opensafely.org).

OpenSAFELY provides a secure software interface allowing the analysis of pseudonymized primary care patient records from England in near real-time within the Electronic Health Records (EHR) vendor’s highly secure data centre, avoiding the need for large volumes of potentially disclosive pseudonymized patient data to be transferred off-site. This, in addition to other technical and organisational controls, minimizes any risk of re-identification.

Similarly pseudonymized datasets from other data providers are securely provided to the EHR vendor and linked to the primary care data. The dataset analysed within OpenSAFELY is based on 24 million people currently registered with GP surgeries using TPP SystmOne software. It includes pseudonymized data such as coded diagnoses, medications and physiological parameters. No free text data are included. Further details on our information governance can be found in the appendix, under <u>information governance and ethics.</u>

### Study Design and Population

We extracted repeated, monthly cohorts of people aged 65 years or older with a known residential status (care home or other) registered with a TPP practice on the 1^st^ of every month from 1^st^ February 2019 until 31^st^ March 2021. Approximately 1% of individuals in TPP do not have a known residential status (6).

### Study Measures

The exposure of interest was residency in a care or nursing home at the 1^st^ of each month. The identification of care homes in OpenSAFELY has been previously described (6). Briefly, individuals’ addresses were matched to the Care Quality Commission (CQC) registry of old-age care homes. Natural language processing was applied to account for spelling inconsistencies, and data cleaning based on the number of residents at a given address was also undertaken. This type of address linkage is thought to have a high positive predictive value (7), however, it is important to note that it cannot identify temporary care home stays and could miss up to a third of permanent care home residents (6). Individuals who were not classified as being a care home resident through this linkage were considered to be living in a private home.

The outcome of interest was mortality; this was ascertained from the ONS. COVID-19 deaths were defined as having an underlying or secondary cause of death listed as COVID-19 (ICD-10 codes U07.1 or U07.2). Specific non-COVID-19 underlying causes of death of interest were also described: deaths due to cancer (ICD-10 chapter code C), cardiovascular disease (ICD-10 chapter I), respiratory disease (ICD-10 chapter J) and dementia (ICD-10 codes F00, F01, F02, F03 and G30). Deaths with any of these underlying causes but a secondary cause of death listed as COVID-19 were considered to be due to COVID-19.

The demographic and clinical characteristics summarised were: age, gender, self-reported ethnicity (5 categories), Nomenclature of Territorial Units for Statistics (NUTS) 1 geographical region of the GP practice, quintile of index of multiple deprivation, stroke, dementia, diabetes, chronic kidney disease, cancer, chronic liver disease, chronic cardiac disease and chronic respiratory disease. Codelists are available through github (https://github.com/opensafely/carehome-noncarehome-death-research/tree/master/codelists). We defined the first pandemic wave as starting on 1^st^ February 2020 and lasting until 31^st^ August 2020, and the second wave as starting on 1^st^ September 2020 until the end of data availability (8).

### Statistical Methods

Mortality proportions were calculated by totalling the number of deaths occurring during a given calendar month among people meeting the inclusion and exclusion criteria at the 1^st^ of that specific month (numerator) and dividing these by the number of individuals meeting the inclusion and exclusion criteria at the beginning of the interval (denominator). These proportions are presented as mortality risks per 100,000 persons per month. Relative risks were calculated by dividing the mortality proportions among care and/or nursing home residents by that among those living in private residences. To account for differences in the age of care home and private home residents, mortality risks for men and women were directly standardised to the European Standard (2013) population using five-year age-bands (9). The directly age-standardised risks (DSR) were calculated by multiplying the mortality rate by the size of each age band in the standard population to generate a total expected number of deaths, and then dividing this by the size of the standard population (10).

Comparative Mortality Figures (CMF) were calculated by taking the DSR among care home residents and dividing these by the DSR among residents of private homes. Confidence intervals for the CMF were calculated following Clayton and Hills, 1993 (11).

Data management was performed using OpenSAFELY tools in Python 3.8 and analyses carried out using R version 3.6.2. All of the code used for data management and analyses, as well as redacted data summaries used for the plots, is available under open licenses for review and re-use at https://github.com/opensafely/carehome-noncarehome-death-research.

### Supplementary Analyses

We conducted several supplementary analyses. Firstly, we described standardised mortality risks of care and nursing home residents separately. Secondly, we stratified the standardised risks by age (above and below 80 years old). Finally, we looked at the characteristics of the population at repeated points in time (1^st^ February 2020 and 1^st^ February 2021).

## Results

### Population Characteristics

There were 4,329,078 individuals aged 65 years or older registered with TPP practices on 1st February 2019, the first cohort extracted, and 95,215 of these (2.2%) were classified as resident in care or nursing homes. The characteristics of these individuals can be seen in Table 1. Residents in care homes compared to residents in private homes were on average older (mean age = 86, SD = 7.83 vs mean age = 75, SD = 7.36), more likely to be female (69.9% vs 53.5%) and much more likely to have underlying comorbidities, including dementia (59.3% vs 3.7%) and a history of stroke (21.9% vs 6.2%).

**Table 1.** Demographic and Clinical Characteristics of Care Home and Private Home Residents on the 1st of February 2019.

|  |  | Overall |  | Care or Nursing Home |  | Private Home |  |
| --- | --- | --- | --- | --- | --- | --- | --- |
|  |  | N | % | N | % | N | % |
| **Total** |  | 4329078 | 100 | 95215 | 100 | 4233863 | 100 |
| Care Home Type | Care Home | 49172 | 1.14 | 49172 | 51.64 |  |  |
|  | Care or Nursing Home | 2364 | 0.05 | 2364 | 2.48 |  |  |
|  | Nursing Home | 43679 | 1.01 | 43679 | 45.87 |  |  |
|  | Private Home | 4233863 | 97.8 |  |  | 4233863 | 100 |
| Gender | Female | 2331960 | 53.87 | 66586 | 69.93 | 2265374 | 53.51 |
|  | Male | 1997118 | 46.13 | 28629 | 30.07 | 1968489 | 46.49 |
| Age in Years | Mean, SD | 75 | 7.54 | 86 | 7.83 | 75 | 7.36 |
| Self - reported Ethnicity | Asian or British Asian | 111123 | 2.57 | 558 | 0.59 | 110565 | 2.61 |
|  | Black | 32845 | 0.76 | 436 | 0.46 | 32409 | 0.77 |
|  | Missing | 1021608 | 23.6 | 25143 | 26.41 | 996465 | 23.54 |
|  | Mixed | 11911 | 0.28 | 187 | 0.2 | 11724 | 0.28 |
|  | Other | 26351 | 0.61 | 322 | 0.34 | 26029 | 0.61 |
|  | White | 3125240 | 72.19 | 68569 | 72.01 | 3056671 | 72.2 |
| Geographical Region | East | 1021027 | 23.59 | 22312 | 23.43 | 998715 | 23.59 |
|  | East Midlands | 766727 | 17.71 | 17091 | 17.95 | 749636 | 17.71 |
|  | London | 165002 | 3.81 | 1914 | 2.01 | 163088 | 3.85 |
|  | North East | 200185 | 4.62 | 4093 | 4.3 | 196092 | 4.63 |
|  | North West | 402111 | 9.29 | 9287 | 9.75 | 392824 | 9.28 |
|  | South East | 318494 | 7.36 | 8179 | 8.59 | 310315 | 7.33 |
|  | South West | 724861 | 16.74 | 15692 | 16.48 | 709169 | 16.75 |
|  | West Midlands | 154434 | 3.57 | 3028 | 3.18 | 151406 | 3.58 |
|  | Yorkshire and The | 575374 | 13.29 | 13607 | 14.29 | 561767 | 13.27 |
|  | Humber |  |  |  |  |  |  |
|  | Missing | 863 | 0.02 | 12 | 0.01 | 851 | 0.02 |
| Quintile of Index of Multiple Deprivation | 1 - Least Deprived | 615577 | 14.22 | 17307 | 18.18 | 598270 | 14.13 |
|  | 2 | 760064 | 17.56 | 19773 | 20.77 | 740291 | 17.49 |
|  | 3 | 974750 | 22.52 | 20814 | 21.86 | 953936 | 22.53 |
|  | 4 | 992850 | 22.93 | 19149 | 20.11 | 973701 | 23 |
|  | 5 - Most Deprived | 967192 | 22.34 | 17715 | 18.61 | 949477 | 22.43 |
|  | Missing | 18645 | 0.43 | 457 | 0.48 | 18188 | 0.43 |
| History of Stroke |  | 288596 | 6.67 | 20885 | 21.93 | 267711 | 6.32 |
| Dementia |  | 211756 | 4.89 | 56417 | 59.25 | 155339 | 3.67 |
| Diabetes |  | 920356 | 21.26 | 22304 | 23.42 | 898052 | 21.21 |
| Chronic Kidney Disease |  | 444229 | 10.26 | 19595 | 20.58 | 424634 | 10.03 |
| Cancer |  | 640900 | 14.8 | 15409 | 16.18 | 625491 | 14.77 |
| Chronic Liver Disease |  | 36287 | 0.84 | 919 | 0.97 | 35368 | 0.84 |
| Chronic Cardiac Disease |  | 864976 | 19.98 | 28901 | 30.35 | 836075 | 19.75 |
| Chronic Respiratory Disease |  | 486564 | 11.24 | 12512 | 13.14 | 474052 | 11.2 |

### Mortality Trends

Age-standardised mortality risks among care home and private home residents according to gender can be seen in Figure 1a – 1c and supplementary materials table S1a-S1c. Among both men and women, all-cause mortality risks increased significantly during the first and second wave of the pandemic, peaking in April 2020. During the peak of the first wave, mortality risks increased to a greater degree in care homes (129% [women] -163% [men] compared to February 2019) compared to private homes (39% [women] - 57% [men] compared February 2019, supplementary materials table S1a). COVID-19 specific mortality followed the same trends, however, there were no marked increases in non-COVID-19 deaths during the second pandemic wave. As expected, the mortality of care home residents was appreciably higher compared to that among private home residents throughout the follow-up period, and the mortality risk for women was lower than that for men in both residential settings.

**Figure 1.**
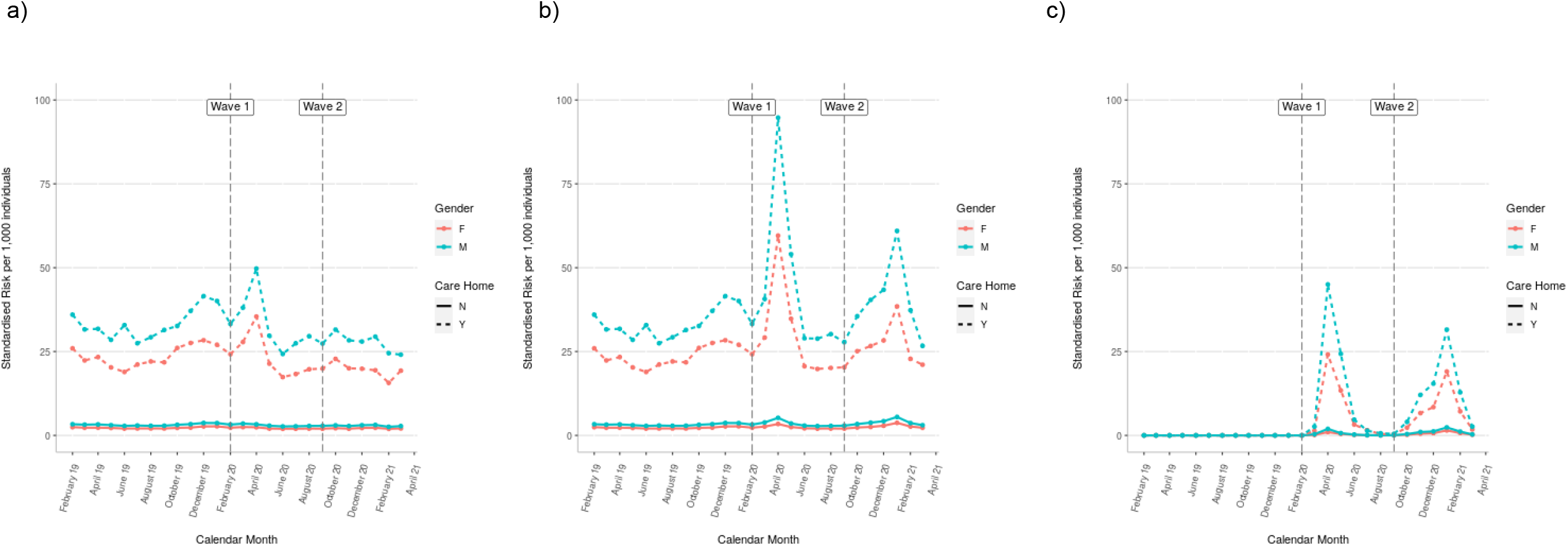
Age-standardised a) All-cause, b) COVID-19, and c) non-COVID-19 Monthly Mortality Risks over Time among Care and Private Home Residents by Gender.

The comparative mortality figures – describing the relative increase in the mortality risk among care home residents compared to private home residents – can be seen in Figure 2a – 2c and supplementary materials table S2a-2c. In February 2019, care home residents had just over 10 times the risk of dying of any cause compared to private home residents (CMF = 10.59, 95%CI = 9.51, 11.81 among women, CMF = 10.82, 95%CI = 9.89, 11.84 among men). By April 2020, this had increased to more than 17 times the mortality risk among residents of private homes of a similar age (CMF = 17.52, 95%CI = 16.38, 18.74 among women, CMF = 18.12, 95%CI = 17.17 – 19.12 among men). By June 2020, the comparative mortality figures had returned to their pre-pandemic value showing approximately a 10-fold increase in the risk of death among care home residents. Of note, the comparative mortality figures did not increase during the second wave, despite a rise in the age-standardised COVID-19 mortality risks among both men and women (CMF at peak [October 2020] = 10.72, 95%CI = 9.67 – 11.89 among women, CMF at peak [January 2021] = 11.09, 95%CI = 10.42-11.81 among men).

**Figure 2.**
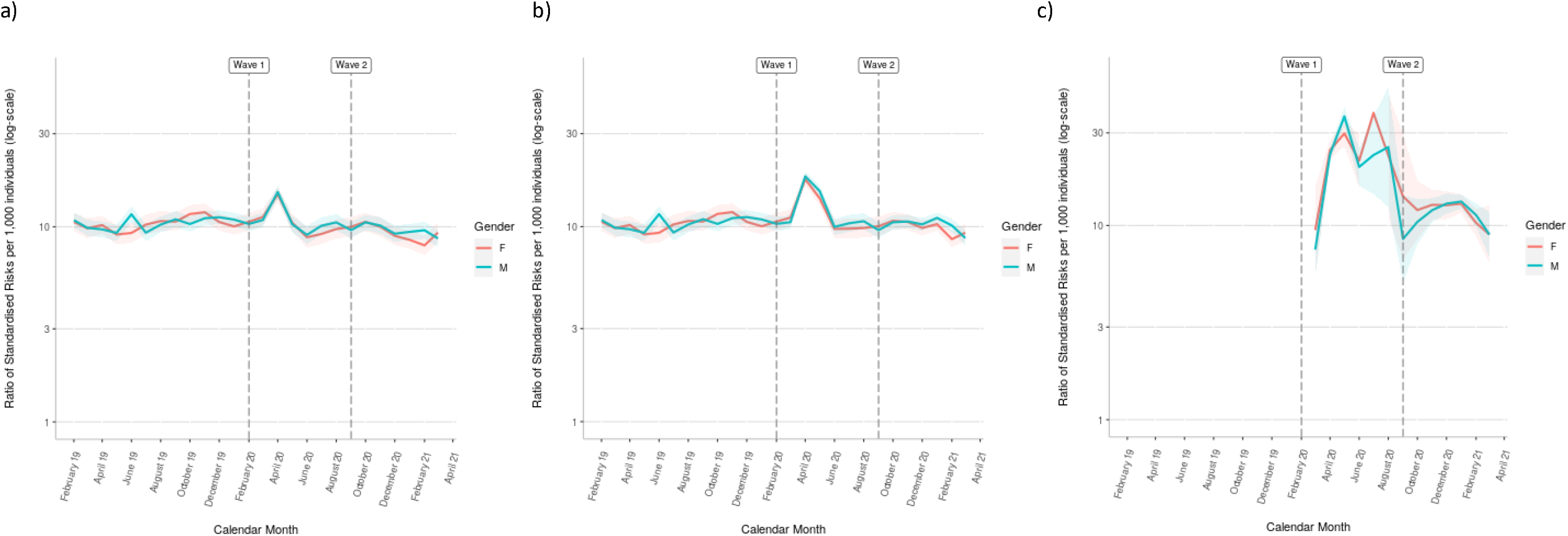
Comparative Mortality Figures (CMFs) Comparing Monthly a) All-cause, b) COVID-19, and c) non-COVID-19 Mortality over Time among Care Home versus Private Home Residents

### Age Differences in Mortality Trends

Crude relative mortality risks stratified by age (Figure 3a-3c) showed a similar overall pattern to the age-standardised mortality risks: a marked peak in the relative risk of death during the first COVID-19 wave, although there was no such increase during the second wave.

**Figure 3.**
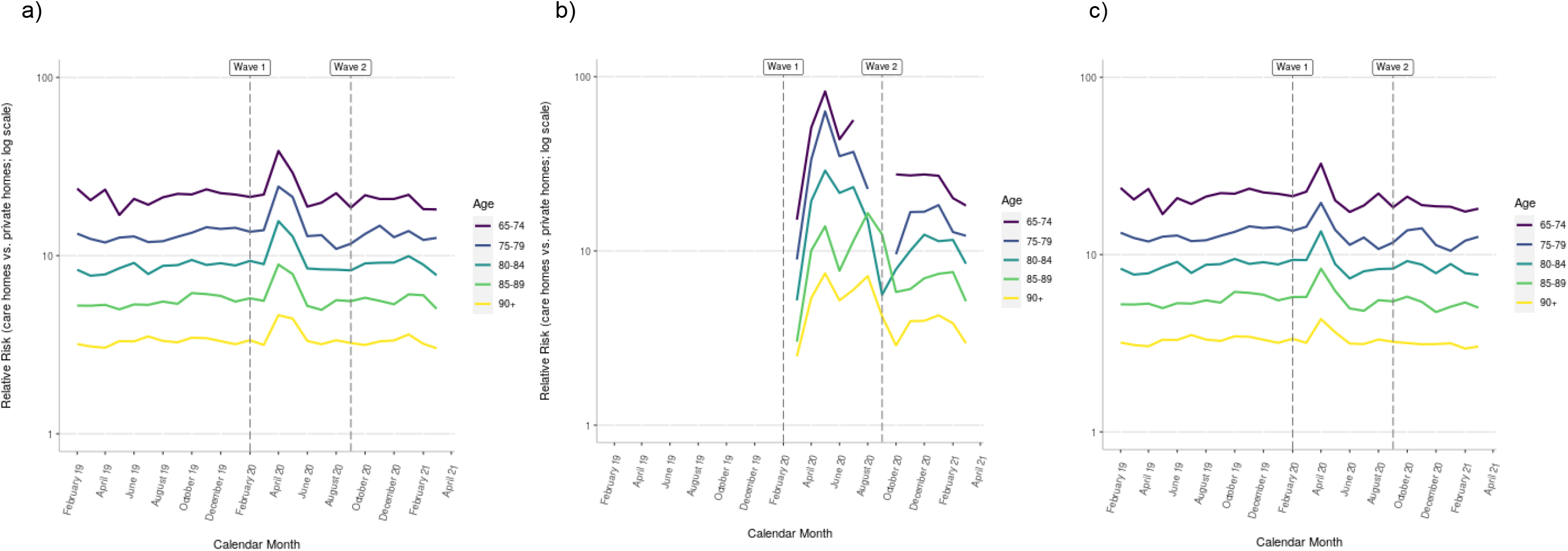
Relative Risk by age of (a) all-cause, (b) COVID-19*, and (c) non-COVID-19 Mortality over Time among Care Home versus Private Home Residents ^*\**^gaps in figure 3b are due to small number redaction, where the numerator values were <=5

The relative risks showed a striking tendency to decline with age, and the largest fluctuations over time were seen in the youngest care home residents. Trends were similar for all-cause, COVID-19 and non-COVID mortality.

### Trends in proportional mortality by cause

Given the increase in non-COVID mortality observed among care home residents during the first pandemic wave, we looked at the proportion of deaths due to key causes of death among care home residents over time (Figure 4a-4b). There were, as expected, a large increase in the proportions of deaths due to COVID-19 during both the first and second wave (Figure 4a). In terms of non-COVID deaths, there was a steady increase in the proportion of deaths due to ‘other’ causes of death over time, and a slight decrease in the proportion of deaths due to respiratory disease (Figure 4b).

**Figure 4.**
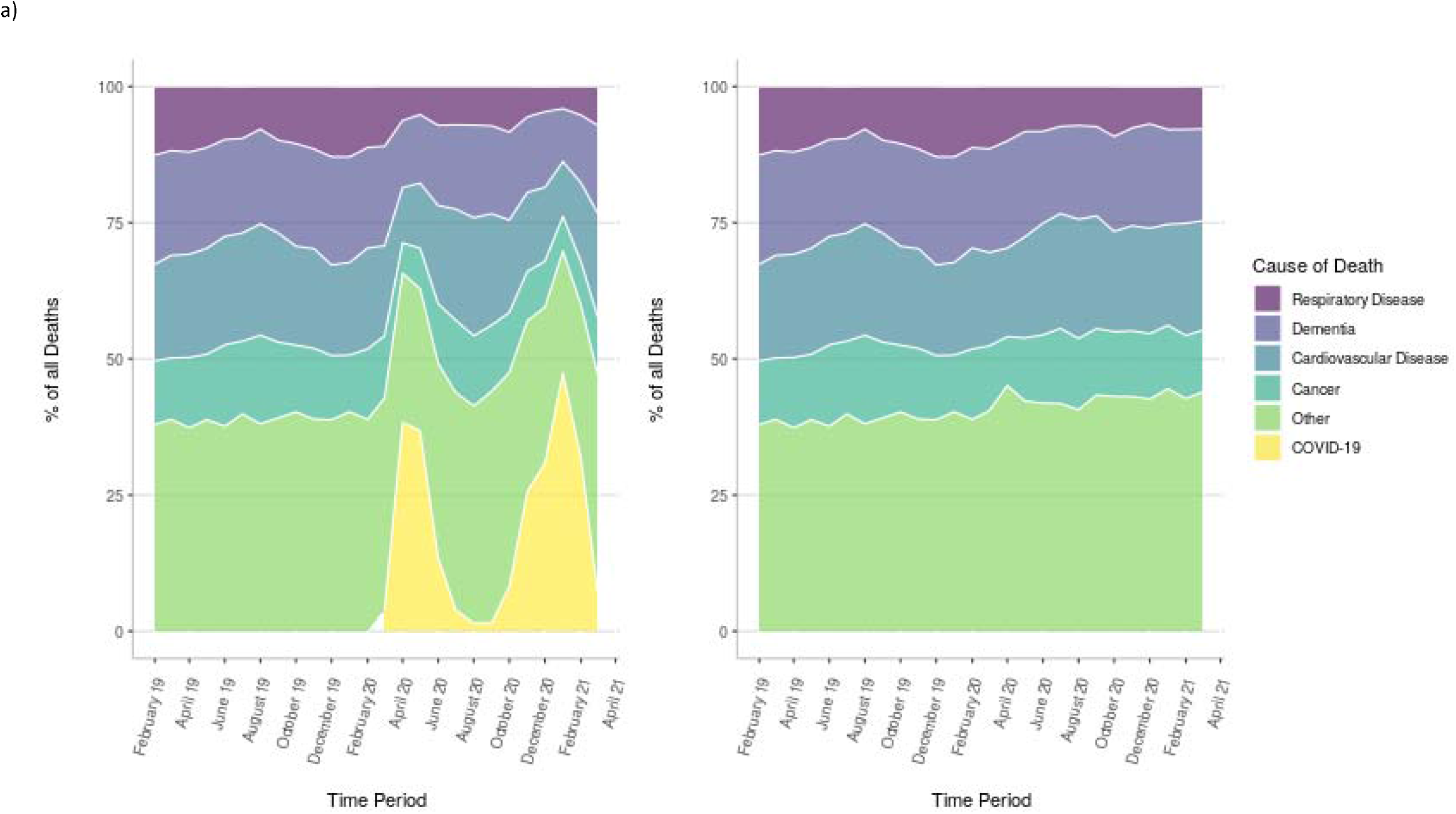
Percentage of Deaths over Time among Residents of Care Homes by cause a) including COVID-19 deaths and b) excluding COVID-19 deaths.

### Supplementary Analyses

A subset of care home residents live in care homes offering nursing care. These individuals tended to have a higher absolute mortality risk compared to those living in care homes where nursing care was not offered (Supplementary Figures 1a-c, 2a-c). Interestingly, although nursing home residents experienced a higher standardised COVID-19 mortality risk than care home residents in the first wave, the COVID-19 mortality risk was almost indistinguishable between these different residential care settings in the second wave (supplementary materials, figures S3a-S3c, S4a-S4c). In terms of age differences, trends described in figure 3a-3c were similar when looking at age-standardised mortality risks stratified by age groups above and below 80 (supplementary figures 3a-3c, 4a-4c). There was little indication that the characteristics of individuals varied over time (supplementary materials, table S3 and S4).

## Discussion

### Summary

People aged 65 years or older living in care homes in England had substantially increased mortality in the first wave of the pandemic compared to those living in private homes. At the height of the first wave, in April 2020, mortality risks increased by 39-57% in private homes compared to 129-163% in care homes compared to the pre-pandemic mortality risks in February 2019. However, during the second wave mortality risks in care and private homes increased to the same proportional degree. This may indicate that improved care home mitigation measures in the second wave reduced the impact of pandemic-related mortality on care home residents. There was no evidence of any substantial shift in the distribution of non-COVID causes of deaths over the time-period studied.

### Comparison to Prior Literature

Hollinghurst et al, using data from Wales, took an approach similar to ours and compared the mortality rate among older care home residents in Wales to that in the general population, and describe findings consistent with ours: an increase in the relative risk of death during 2020 among care home residents compared to residents in private home (12). They adjusted for a wide range of comorbidities and reported hazard ratios increasing from 2.15 (95%CI = 2.11 – 2.20) to 2.94 (95%CI = 2.81 – 3.08) among care home residents. The difference in the size of the relative mortality in this study, compared to the age-standardised mortality risks we describe, highlight the importance of interpreting relative risks with close attention to the specific adjustment sets used in each study.

Several studies have described a large excess mortality within care homes in the UK during the pandemic, largely restricted to care homes with recorded COVID-19 outbreaks (4,5).

This indicates that this excess mortality has largely been due to COVID-19, irrespective of the cause of death registered on death certificates. Our analyses support the conclusion; during the first pandemic wave a marked increase in non-COVID-19 deaths was observed - no such increase was present during the second wave. Had the excess non-COVID deaths during the first pandemic wave been due to changes in the care provided to care home residents as a result of staff being stretched due to increased sick leave, or hospitals being overwhelmed, we would have expected similar increases during the second wave as well.

Evidence from the pre-pandemic period in the UK have also found a substantially higher mortality rate among care home residents compared to private home residents (13,14). Shah et al, used data from CPRD to describe the mortality experience of care home residents in the UK, and report an approximate four-fold increase in the risk of death (13). A more recent analysis of Pujol et al, also using CPRD data, showed that this has increased considerably between 2006 and 2015 (14). Interestingly, they also show a considerably higher relative risk of death for younger care home residents compared to private home residents. Such an age difference could reflect the increased frailty of individuals who need to access care at relatively young ages. In general, our findings are consistent with those described by Pujol et al, despite covering a more recent calendar year period.

### Strengths and Limitations

Our study has several strengths. Most notably, we had access to clearly defined and time-updated population at risk (denominators) as well as deaths (numerators) by place of residence through the address linkage to CQC care home addresses in OpenSAFELY-TPP. This has allowed us to contrast the absolute mortality risks of care home residents to those in private homes, and therefore builds on updates provided by the ONS on place of death, which only provides numerator data.

The most significant limitation concerns the identification of care home residents in UK EHR data (6). Here, we used an address linkage with CQC data. Although this likely has a high positive predictive value (7), the prevalence of care home residency is approximately a third lower than indicated based on estimates from the 2011 census (15). Location of death should not have impacted on these analyses, as the ONS data captures deaths occurring both in hospital and within the community. However, we cannot rule out that some of the differences we observe in the mortality risks between care home and private home residents might be due to data quality issues, for example if there has been poorer ascertainment of whether someone lives in a care home during the second wave due to more dynamic movements of people. It should also be noted that our findings may not be generalisable to the English total care home population, as TPP only covers a proportion of English residents.

### Interpretation and Policy Implications

In contrast to the first wave, the rise in mortality risks during the second pandemic wave affected private and care home residents to the same proportional degree. Absolute mortality risks among care home residents were also lower compared to the first peak. It is possible that although preventative measures may not have been sufficient to stop the introduction of infections into care homes during the second wave, they had an impact on spread within care homes following the identification of the first case – thus preventing unnoticed or uncontrolled outbreaks. There may also have been an element of short-term mortality displacement, so-called ‘harvesting’ (16), with the population of care home residents during the second wave representing a ‘survivor’ population. To investigate this possibility, we conducted a post-hoc analysis describing the characteristics of care home residents over time (supplementary materials table S3-S4); however, this did not indicate large changes in the age or prevalence of underlying comorbidities. Nonetheless, some element of mortality displacement is likely, as we would expect residents who died during the first wave to have been replaced by generally younger, fitter residents with presumably a lower mortality. Pre-existing immunity among survivors of the first wave may also have contributed to slowing the spread of the infection during the second wave. The VIVALDI study of 100 long-term care facilities in England found an antibody prevalence as high as 33% among residents in June 2020, with the presence of IgG antibodies strongly reducing the risk of reinfection among residents during the second wave (HR = 0.39, 95%CI = 0.19 – 0.77) (17). The UK vaccination program may also have contributed in part, although the mortality peak in January 2021 occurred before all care home residents in the UK had been vaccinated (18). Due to the time taken for immunity to develop, factors in addition to the vaccination programme are likely to have played a role. Nevertheless, it is not in doubt that the UK’s rapid vaccination roll out has markedly reduced the risk of becoming severely ill or dying due to COVID-19 for older adults, including care home residents (19).

### Further Research

Our data presents several questions for future research. Firstly, although analyses of factors associated with outbreaks in care homes have been done during the first wave; extending these analyses to study variation over time would be of great interest. Secondly, analyses studying the impact of the vaccination program among care home residents would be valuable. Finally, although the identification of care home residents through an address linkage is a strength, we are also aware that this will misclassify some individuals as residents in private homes (6). As others have argued (20,21), we strongly believe that the development of data infrastructure that can identify spells of care home residence should be a priority in order to allow the healthcare needs of this vulnerable population to be comprehensively characterised.

## Conclusion

Our analysis highlights the stark impact of the COVID-19 pandemic on care homes in England, with residents suffering a disproportionately increased mortality risk during the first wave compared to individuals of a similar age living in private residences. Although absolute mortality risks increased during the second wave, these remained below the first wave peak, and the relative mortality risk of care home residents compared to individuals living in private residences remained unchanged – possibly reflecting preventative measures such as increased testing and infection control measures within care homes, or high levels of pre-existing immunity. Taken together, our data highlights the extreme susceptibility of care home residents to COVID-19 and reaffirms the importance of targeting protective measures towards those living in residential care.

## Data Availability

For security and privacy reasons, OpenSAFELY is very different to other approaches for EHR data analysis. The platform does not give researchers unconstrained access to view large volumes of pseudonymised and disclosive patient data, either via download or via a remote desktop. Instead we have produced a series of open source tools that enable researchers to use flexible, pragmatic, but standardised approaches to process raw electronic health records data into 'research ready' datasets, and to check that this has been done correctly, without needing to access the patient data directly. Using this data management framework we also generate bespoke dummy datasets. These dummy datasets are used by researchers to develop analysis code in the open, using GitHub. When their data management and data analysis scripts are capable of running to completion, and passing all tests in the OpenSAFELY framework, they are finally sent through to be executed against the real data inside the secure environment, using the OpenSAFELY jobs runner, inside a container using Docker, without the researcher needing access to that raw potential disclosive pseudonymised data themselves. The non-disclosive summary results output tables, logs, and graphs are then manually reviewed, as in other systems, before release.As part of building that resource for the community, over the next six months we are working with NHS England to cautiously on-board a small number of external pilot users to develop their analyses on OpenSAFELY. This process is described in further detail on our webpage, here: https://opensafely.org/onboarding-new-users/.

https://github.com/opensafely/carehome-noncarehome-death-research

## Administrative

## Acknowledgements

We are very grateful for all the support received from the TPP Technical Operations team throughout this work, and for generous assistance from the information governance and database teams at NHS England / NHSX.

## Funding

This work was supported by the Medical Research Council MR/V015737/1. TPP provided technical expertise and infrastructure within their data centre *pro bono* in the context of a national emergency.

BG’s work on better use of data in healthcare more broadly is currently funded in part by: NIHR Oxford Biomedical Research Centre, NIHR Applied Research Collaboration Oxford and Thames Valley, the Mohn-Westlake Foundation, NHS England, and the Health Foundation; all DataLab staff are supported by BG’s grants on this work. LS reports grants from Wellcome, MRC, NIHR, UKRI, British Council, GSK, British Heart Foundation, and Diabetes UK outside this work. AS and JT are employed by LSHTM on fellowships sponsored by GSK. KB holds a Sir Henry Dale fellowship jointly funded by Wellcome and the Royal Society (107731/Z/15/Z). HIM is funded by the National Institute for Health Research (NIHR) Health Protection Research Unit in Immunisation, a partnership between Public Health England and LSHTM. AYSW holds a fellowship from BHF. ID holds grants from NIHR and GSK. RM holds a Sir Henry Wellcome Fellowship funded by the Wellcome Trust (201375/Z/16/Z). HF holds a UKRI fellowship.

The views expressed are those of the authors and not necessarily those of the NIHR, NHS England, Public Health England or the Department of Health and Social Care.

Funders had no role in the study design, collection, analysis, and interpretation of data; in the writing of the report; and in the decision to submit the article for publication.

### Information governance and ethical approval

NHS England is the data controller; TPP is the data processor; and the key researchers on OpenSAFELY are acting on behalf of NHS England. This implementation of OpenSAFELY is hosted within the TPP environment which is accredited to the ISO 27001 information security standard and is NHS IG Toolkit compliant;[1,2] patient data has been pseudonymised for analysis and linkage using industry standard cryptographic hashing techniques; all pseudonymised datasets transmitted for linkage onto OpenSAFELY are encrypted; access to the platform is via a virtual private network (VPN) connection, restricted to a small group of researchers; the researchers hold contracts with NHS England and only access the platform to initiate database queries and statistical models; all database activity is logged; only aggregate statistical outputs leave the platform environment following best practice for anonymisation of results such as statistical disclosure control for low cell counts.[3] The OpenSAFELY research platform adheres to the obligations of the UK General Data Protection Regulation (GDPR) and the Data Protection Act 2018. In March 2020, the Secretary of State for Health and Social Care used powers under the UK Health Service (Control of Patient Information) Regulations 2002 (COPI) to require organisations to process confidential patient information for the purposes of protecting public health, providing healthcare services to the public and monitoring and managing the COVID-19 outbreak and incidents of exposure; this sets aside the requirement for patient consent.[4] Taken together, these provide the legal bases to link patient datasets on the OpenSAFELY platform. GP practices, from which the primary care data are obtained, are required to share relevant health information to support the public health response to the pandemic, and have been informed of the OpenSAFELY analytics platform.

This study was approved by the Health Research Authority (REC reference 20/LO/0651) and by the LSHTM Ethics Board (reference 21863).

### Guarantor

BG/LS is guarantor.

## Research In Context

### Evidence before this study

Residents of care homes in the UK and elsewhere are known to have been severely affected by the COVID-19 pandemic. In the UK this has been clearly demonstrated by very large increases in the number of excess deaths occurring in care homes in first and second waves 2020/21, and by detailed studies in Scotland and Wales up to the summer of 2020. However, to date there have not been any large-scale studies of care home mortality in England over the first two pandemic waves that have been based on follow-up of care home residents regardless of whether they died where they lived or in hospital.

### Added value of the study

Much of previously published literature on COVID-19 in care homes have focused on excess mortality. Our study uses an address linkage to define a population of care home residents and quantifies their mortality risk compared to individuals of a similar age residents in private homes between February 2019 and March 2021. We find that the first COVID-19 wave in the UK has had a disproportionate impact on care home residents. Age-standardised mortality risks were approximately 10-fold higher for care home residents compared to private home residents in the pre-pandemic period; this increased to approximately 17-fold during the peak of the first pandemic wave. However, during the second wave, mortality risks increased to the same proportional degree among care home residents and residents of private homes.

### Implications of all of the available evidence

Despite UK governmental policy aimed at protecting care homes, residents in England experienced disproportionately high mortality during the first COVID-19 pandemic wave. It is possible that some degree of immunity induced by infections in the first wave, improved protective measures or changes in the underlying frailty of the populations studied may have contributed to the absence of such an impact during the second wave. Our data supports targeting protective measures, including vaccinations, towards residents as well as ensuring social care staff have the resources required to implement infection control measures.

## References

1. Mortality associated with COVID-19 outbreaks in care homes: early international evidence [Internet]. Resources to support community and institutional Long-Term Care responses to COVID-19. 2020 [cited 2021 Jun 23]. Available from: https://ltccovid.org/2020/04/12/mortality-associated-with-covid-19-outbreaks-in-care-homes-early-international-evidence/

2. Deaths involving COVID-19 in the care sector, England and Wales - Office for National Statistics [Internet]. [cited 2021 Apr 8]. Available from: https://www.ons.gov.uk/peoplepopulationandcommunity/birthsdeathsandmarriages/deat hs/articles/deathsinvolvingcovid19inthecaresectorenglandandwales/deathsoccurringupto 12june2020andregisteredupto20june2020provisional

3. Hodgson H, Grimm F, Vestesson E, Brine R,, Deeny S. Adult social care and COVID-19: Assessing the impact on social care users and staff in England so far | The Health Foundation [Internet]. 2020 [cited 2021 Apr 8]. Available from: https://doi.org/10.37829/HF-2020-Q16

4. Burton JK, Bayne G, Evans C, Garbe F, Gorman D, Honhold N, et al. Evolution and effects of COVID-19 outbreaks in care homes: a population analysis in 189 care homes in one geographical region of the UK. Lancet Healthy Longev. 2020 Oct 1;1(1):e21–31.

5. Morciano M, Stokes J, Kontopantelis E, Hall I, Turner AJ. Excess mortality for care home residents during the first 23 weeks of the COVID-19 pandemic in England: a national cohort study. BMC Med. 2021 Mar 5;19(1):71.

6. Schultze A, Bates C, Cockburn J, MacKenna B, Nightingale E, Curtis H, et al. Identifying Care Home Residents in Electronic Health Records - An OpenSAFELY Short Data Report. Wellcome Open Res. 2021 Apr 27;6:90.

7. Burton JK, Marwick CA, Galloway J, Hall C, Nind T, Reynish EL, et al. Identifying care-home residents in routine healthcare datasets: a diagnostic test accuracy study of five methods. Age Ageing. 2019 Jan 1;48(1):114–21.

8. Mathur R, Rentsch CT, Morton CE, Hulme WJ, Schultze A, MacKenna B, et al. Ethnic differences in SARS-CoV-2 infection and COVID-19-related hospitalisation, intensive care unit admission, and death in 17 million adults in England: an observational cohort study using the OpenSAFELY platform. The Lancet. 2021 May 8;397(10286):1711–24.

9. Revision of the European Standard Population - Report of Eurostat’s task force - 2013 edition [Internet]. [cited 2021 Apr 8]. Available from: https://ec.europa.eu/eurostat/web/products-manuals-and-guidelines/-/KS-RA-13-028

10. Kirkwood B, Sterne J. Essential Medical Statistics, 2nd Edition | Wiley. 2003.

11. Clayton D, Hills M. Statistical Models in Epidemiology. Reprint edition. OUP Oxford; 1993. 373 p.

12. Hollinghurst J, Lyons J, Fry R, Akbari A, Gravenor M, Watkins A, et al. The impact of COVID-19 on adjusted mortality risk in care homes for older adults in Wales, UK: a retrospective population-based cohort study for mortality in 2016–2020. Age Ageing. 2021 Jan 1;50(1):25–31.

13. Shah SM, Carey IM, Harris T, DeWilde S, Cook DG. Mortality in older care home residents in England and Wales. Age Ageing. 2013 Mar 1;42(2):209–15.

14. Pujol FE, Hancock R, Morciano M. Trends in survival of older care home residents in England: a 10-year multi-cohort study. Soc Sci Med. 2021 Mar 31;113883.

15. Changes in the Older Resident Care Home Population between 2001 and 2011 - Office for National Statistics [Internet]. [cited 2021 Apr 16]. Available from: https://www.ons.gov.uk/peoplepopulationandcommunity/birthsdeathsandmarriages/ageing/articles/changesintheolderresidentcarehomepopulationbetween2001and2011/2014-08-01

16. Alicandro G, Remuzzi G, Vecchia CL. Italy’s first wave of the COVID-19 pandemic has ended: no excess mortality in May, 2020. The Lancet. 2020 Sep 12;396(10253):e27–8.

17. Krutikov M, Palmer T, Tut G, Fuller C, Shrotri M, Williams H, et al. Incidence of SARS-CoV-2 infection according to baseline antibody status in staff and residents of 100 Long Term Care Facilities (VIVALDI study). medRxiv. 2021 Mar 10;2021.03.08.21253110.

18. NHS EnglandL» NHS confirms COVID jab now offered at every eligible care home in England [Internet]. [cited 2021 Apr 22]. Available from: https://www.england.nhs.uk/2021/02/nhs-confirms-covid-jab-now-offered-at-every-eligible-care-home-in-england/

19. Bernal JL, Andrews N, Gower C, Stowe J, Robertson C, Tessier E, et al. Early effectiveness of COVID-19 vaccination with BNT162b2 mRNA vaccine and ChAdOx1 adenovirus vector vaccine on symptomatic disease, hospitalisations and mortality in older adults in England. medRxiv. 2021 Mar 2;2021.03.01.21252652.

20. Burton JK, Goodman C, Guthrie B, Gordon AL, Hanratty B, Quinn TJ. Closing the UK care home data gap – methodological challenges and solutions: Closing the UK care home data gap. Int J Popul Data Sci [Internet]. 2020 Dec 15 [cited 2021 Apr 8];5(4). Available from: https://ijpds.org/article/view/1391

21. Hanratty B, Burton JK, Goodman C, Gordon AL, Spilsbury K. Covid-19 and lack of linked datasets for care homes. BMJ. 2020 Jun 24;369:m2463.

