## supplemental materials for "Mortality among Care Home Residents in England during the first and second waves of the COVID-19 pandemic: an analysis of 4.3 million adults over the age of 65"

### Supplementary Data

Note: key tables referenced in text are provided in the supplementary data, however, all raw output is also available on github: <https://github.com/opensafely/carehome-noncarehome-death-research/tree/master/released_outputs/output/tables>

#### **Figure S1a-c. Age-standardised a) all-cause, b) covid and c) non-covid mortality risks according to care home type, among men.**

a) b)	c)

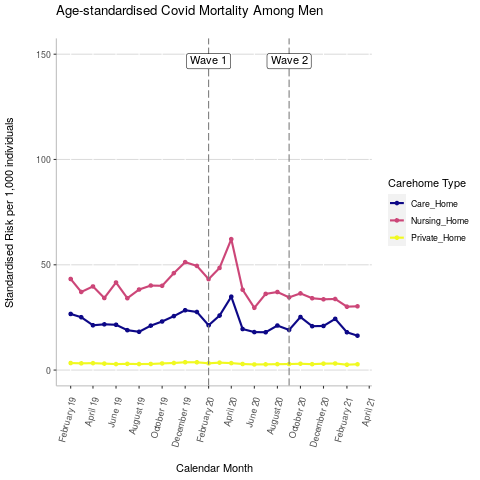

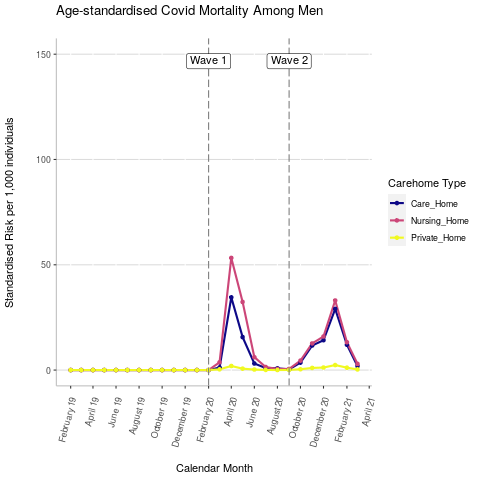

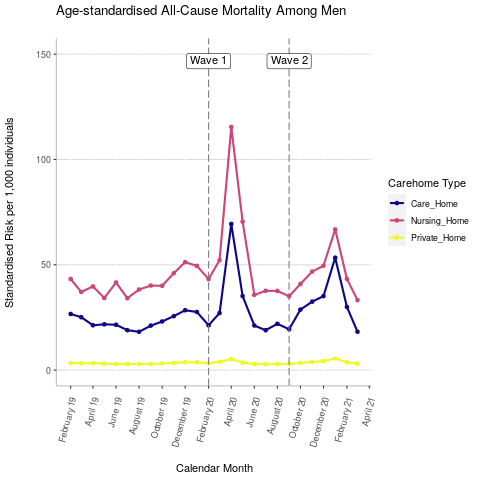

#### **Figure S2a-c Age-standardised risks according to care home type, among women.**

a) b)	c)

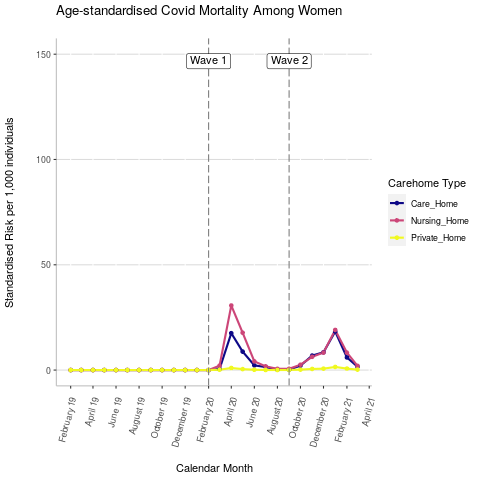

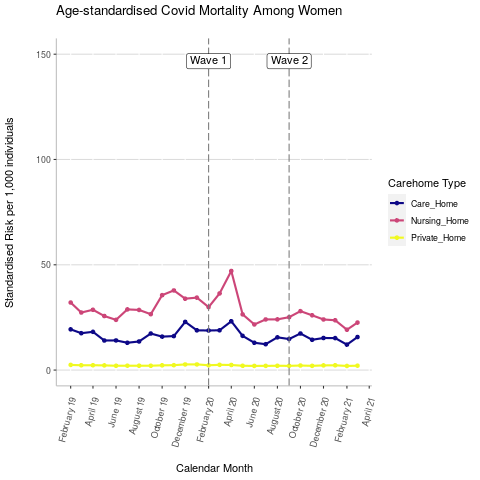

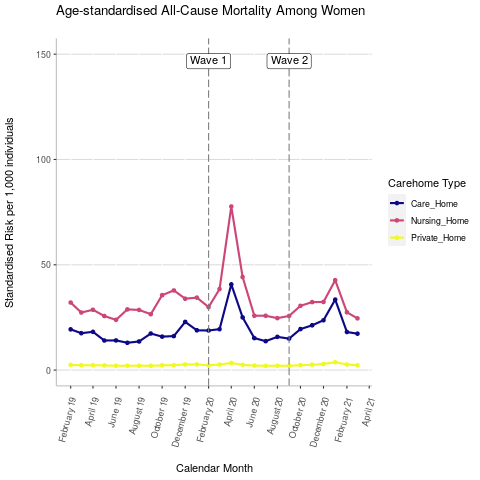

#### **Figure S3a-c. Age-standardised relative a) all-cause, b) covid and c) non-covid mortality risks according to age group, among men**

a) b)	c)

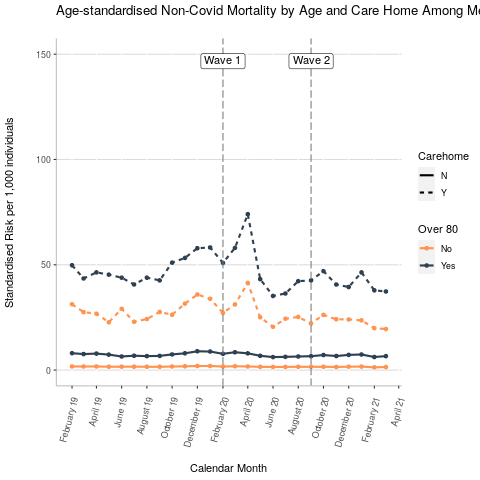

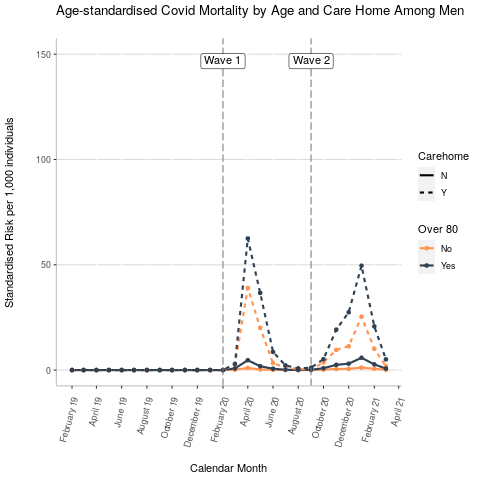

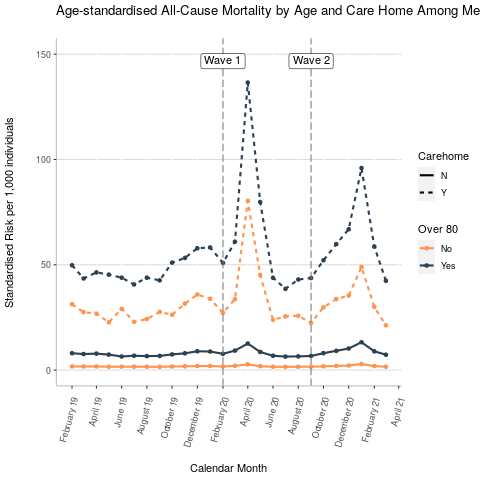

#### **Figure S4a-c. Age-standardised relative a) all-cause, b) covid and c) non-covid mortality risks according to age group, among women**

a) b)	c)

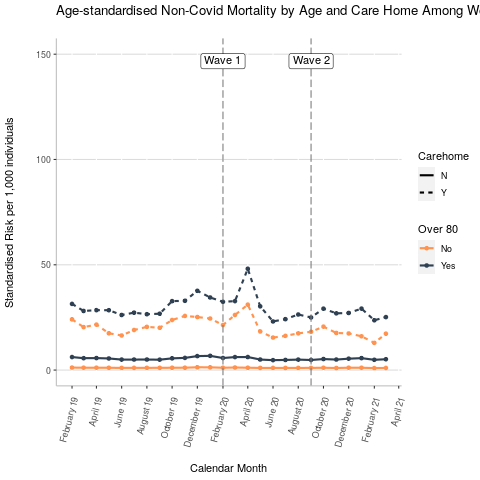

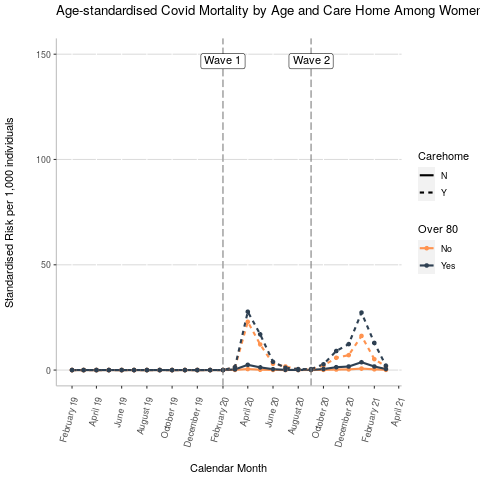

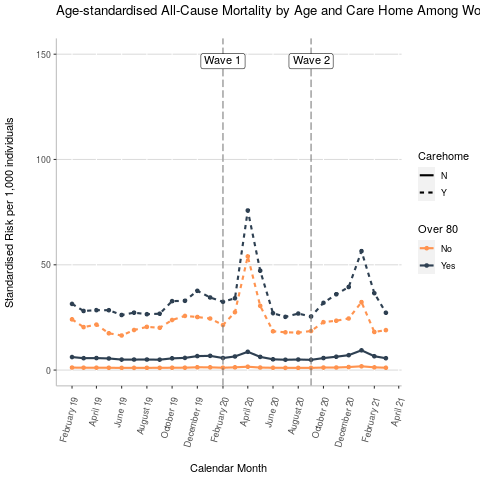

#### **Table S1a. Age-standardised all-cause mortality risks by gender**

| **Gender** | **Date** | **Care or Nursing Home Standardised Risk** | **Care or Nursing Home Confidence Interval** | **Private Home Standardised Risk** | **Private Home Confidence Interval** |
| --- | --- | --- | --- | --- | --- |
| F | 01/02/2019 | 25.96 | 23.26-28.66 | 2.45 | 2.39-2.51 |
| F | 01/03/2019 | 22.35 | 19.82-24.89 | 2.27 | 2.21-2.34 |
| F | 01/04/2019 | 23.34 | 20.92-25.76 | 2.29 | 2.23-2.35 |
| F | 01/05/2019 | 20.27 | 18.07-22.47 | 2.22 | 2.16-2.28 |
| F | 01/06/2019 | 18.92 | 16.75-21.08 | 2.03 | 1.98-2.09 |
| F | 01/07/2019 | 21.13 | 18.79-23.46 | 2.06 | 2-2.12 |
| F | 01/08/2019 | 22.05 | 19.53-24.57 | 2.06 | 2-2.12 |
| F | 01/09/2019 | 21.79 | 19.43-24.14 | 2.05 | 1.99-2.11 |
| F | 01/10/2019 | 26.07 | 23.46-28.68 | 2.24 | 2.18-2.3 |
| F | 01/11/2019 | 27.55 | 24.82-30.27 | 2.32 | 2.26-2.38 |
| F | 01/12/2019 | 28.37 | 25.72-31.03 | 2.68 | 2.62-2.75 |
| F | 01/01/2020 | 27.02 | 24.46-29.58 | 2.69 | 2.62-2.76 |
| F | 01/02/2020 | 24.22 | 21.79-26.64 | 2.28 | 2.22-2.34 |
| F | 01/03/2020 | 29.17 | 26.38-31.96 | 2.62 | 2.56-2.69 |
| F | 01/04/2020 | 59.57 | 55.88-63.27 | 3.4 | 3.33-3.47 |
| F | 01/05/2020 | 34.77 | 31.96-37.58 | 2.49 | 2.42-2.55 |
| F | 01/06/2020 | 20.61 | 18.39-22.82 | 2.12 | 2.06-2.17 |
| F | 01/07/2020 | 19.81 | 17.57-22.05 | 2.03 | 1.97-2.09 |
| F | 01/08/2020 | 20.11 | 17.96-22.26 | 2.04 | 1.99-2.1 |
| F | 01/09/2020 | 20.3 | 18.04-22.57 | 2.02 | 1.96-2.08 |
| F | 01/10/2020 | 25.1 | 22.62-27.58 | 2.34 | 2.28-2.4 |
| F | 01/11/2020 | 26.65 | 24.24-29.07 | 2.52 | 2.46-2.58 |
| F | 01/12/2020 | 28.31 | 25.84-30.78 | 2.87 | 2.81-2.94 |
| F | 01/01/2021 | 38.46 | 35.63-41.28 | 3.73 | 3.66-3.81 |
| F | 01/02/2021 | 22.81 | 20.76-24.86 | 2.64 | 2.58-2.71 |
| F | 01/03/2021 | 21.08 | 18.92-23.25 | 2.26 | 2.2-2.32 |
| M | 01/02/2019 | 36 | 32.94-39.06 | 3.33 | 3.24-3.41 |
| M | 01/03/2019 | 31.6 | 28.76-34.44 | 3.2 | 3.11-3.28 |
| M | 01/04/2019 | 31.77 | 28.99-34.55 | 3.27 | 3.19-3.36 |
| M | 01/05/2019 | 28.49 | 25.86-31.11 | 3.06 | 2.98-3.14 |
| M | 01/06/2019 | 32.87 | 29.93-35.81 | 2.84 | 2.76-2.91 |
| M | 01/07/2019 | 27.47 | 24.91-30.04 | 2.94 | 2.86-3.02 |
| M | 01/08/2019 | 29.27 | 26.66-31.87 | 2.85 | 2.77-2.93 |
| M | 01/09/2019 | 31.44 | 28.53-34.34 | 2.88 | 2.8-2.96 |
| M | 01/10/2019 | 32.62 | 29.89-35.34 | 3.16 | 3.07-3.24 |
| M | 01/11/2019 | 37.11 | 34.11-40.11 | 3.35 | 3.27-3.44 |
| M | 01/12/2019 | 41.51 | 38.31-44.71 | 3.71 | 3.62-3.8 |
| M | 01/01/2020 | 40.09 | 37.08-43.11 | 3.68 | 3.59-3.77 |
| M | 01/02/2020 | 33.23 | 30.5-35.96 | 3.21 | 3.13-3.3 |
| M | 01/03/2020 | 40.66 | 37.62-43.71 | 3.86 | 3.77-3.95 |
| M | 01/04/2020 | 94.76 | 90.25-99.26 | 5.23 | 5.12-5.34 |
| M | 01/05/2020 | 54.02 | 50.53-57.51 | 3.55 | 3.46-3.63 |
| M | 01/06/2020 | 28.96 | 26.33-31.59 | 2.9 | 2.82-2.98 |
| M | 01/07/2020 | 28.84 | 26.14-31.54 | 2.77 | 2.69-2.85 |
| M | 01/08/2020 | 30.2 | 27.47-32.93 | 2.83 | 2.75-2.91 |
| M | 01/09/2020 | 27.8 | 25.3-30.31 | 2.9 | 2.82-2.97 |
| M | 01/10/2020 | 35.54 | 32.73-38.36 | 3.37 | 3.29-3.46 |
| M | 01/11/2020 | 40.4 | 37.35-43.45 | 3.8 | 3.71-3.89 |
| M | 01/12/2020 | 43.41 | 40.33-46.5 | 4.23 | 4.13-4.32 |
| M | 01/01/2021 | 60.99 | 57.45-64.53 | 5.5 | 5.39-5.61 |
| M | 01/02/2021 | 37.32 | 34.44-40.2 | 3.68 | 3.59-3.77 |
| M | 01/03/2021 | 26.67 | 24.33-29.02 | 3.05 | 2.97-3.13 |

#### **Table S1b. Age-standardised COVID mortality risks by gender**

| **Gender** | **Date** | **Care or Nursing Home Standardised Risk** | **Care or Nursing Home Confidence Interval** | **Private Home Standardised Risk** | **Private Home Confidence Interval** |
| --- | --- | --- | --- | --- | --- |
| F | 01/02/2019 | 0 | 0-0 | 0 | 0-0 |
| F | 01/03/2019 | 0 | 0-0 | 0 | 0-0 |
| F | 01/04/2019 | 0 | 0-0 | 0 | 0-0 |
| F | 01/05/2019 | 0 | 0-0 | 0 | 0-0 |
| F | 01/06/2019 | 0 | 0-0 | 0 | 0-0 |
| F | 01/07/2019 | 0 | 0-0 | 0 | 0-0 |
| F | 01/08/2019 | 0 | 0-0 | 0 | 0-0 |
| F | 01/09/2019 | 0 | 0-0 | 0 | 0-0 |
| F | 01/10/2019 | 0 | 0-0 | 0 | 0-0 |
| F | 01/11/2019 | 0 | 0-0 | 0 | 0-0 |
| F | 01/12/2019 | 0 | 0-0 | 0 | 0-0 |
| F | 01/01/2020 | 0 | 0-0 | 0 | 0-0 |
| F | 01/02/2020 | 0 | 0-0 | 0 | 0-0 |
| F | 01/03/2020 | 1.33 | 0.64-2.02 | 0.14 | 0.13-0.16 |
| F | 01/04/2020 | 24.13 | 21.67-26.6 | 0.99 | 0.95-1.03 |
| F | 01/05/2020 | 13.39 | 11.57-15.21 | 0.45 | 0.42-0.48 |
| F | 01/06/2020 | 3.22 | 2.26-4.19 | 0.15 | 0.13-0.17 |
| F | 01/07/2020 | 1.54 | 0.87-2.21 | 0.04 | 0.03-0.05 |
| F | 01/08/2020 | 0.39 | 0.13-0.64 | 0.02 | 0.01-0.02 |
| F | 01/09/2020 | 0.39 | 0.11-0.66 | 0.03 | 0.02-0.03 |
| F | 01/10/2020 | 2.29 | 1.53-3.06 | 0.19 | 0.17-0.21 |
| F | 01/11/2020 | 6.65 | 5.52-7.78 | 0.52 | 0.49-0.55 |
| F | 01/12/2020 | 8.43 | 7.09-9.76 | 0.66 | 0.63-0.69 |
| F | 01/01/2021 | 19.05 | 17.02-21.08 | 1.47 | 1.42-1.52 |
| F | 01/02/2021 | 7.16 | 6.05-8.26 | 0.69 | 0.66-0.73 |
| F | 01/03/2021 | 1.78 | 1.19-2.38 | 0.2 | 0.18-0.22 |
| M | 01/02/2019 | 0 | 0-0 | 0 | 0-0 |
| M | 01/03/2019 | 0 | 0-0 | 0 | 0-0 |
| M | 01/04/2019 | 0 | 0-0 | 0 | 0-0 |
| M | 01/05/2019 | 0 | 0-0 | 0 | 0-0 |
| M | 01/06/2019 | 0 | 0-0 | 0 | 0-0 |
| M | 01/07/2019 | 0 | 0-0 | 0 | 0-0 |
| M | 01/08/2019 | 0 | 0-0 | 0 | 0-0 |
| M | 01/09/2019 | 0 | 0-0 | 0 | 0-0 |
| M | 01/10/2019 | 0 | 0-0 | 0 | 0-0 |
| M | 01/11/2019 | 0 | 0-0 | 0 | 0-0 |
| M | 01/12/2019 | 0 | 0-0 | 0 | 0-0 |
| M | 01/01/2020 | 0 | 0-0 | 0 | 0-0 |
| M | 01/02/2020 | 0 | 0-0 | 0 | 0-0 |
| M | 01/03/2020 | 2.6 | 1.76-3.44 | 0.35 | 0.32-0.37 |
| M | 01/04/2020 | 45.01 | 41.83-48.2 | 1.93 | 1.87-2 |
| M | 01/05/2020 | 24.31 | 22.01-26.6 | 0.67 | 0.63-0.71 |
| M | 01/06/2020 | 4.68 | 3.73-5.63 | 0.23 | 0.21-0.26 |
| M | 01/07/2020 | 1.39 | 0.8-1.98 | 0.06 | 0.05-0.07 |
| M | 01/08/2020 | 0.63 | 0.22-1.03 | 0.02 | 0.02-0.03 |
| M | 01/09/2020 | 0.43 | 0.22-0.63 | 0.05 | 0.04-0.06 |
| M | 01/10/2020 | 4.01 | 2.98-5.05 | 0.38 | 0.36-0.41 |
| M | 01/11/2020 | 12.06 | 10.43-13.68 | 1.01 | 0.96-1.05 |
| M | 01/12/2020 | 15.44 | 13.76-17.11 | 1.19 | 1.14-1.24 |
| M | 01/01/2021 | 31.55 | 29.04-34.06 | 2.37 | 2.3-2.45 |
| M | 01/02/2021 | 12.83 | 11.16-14.49 | 1.13 | 1.08-1.18 |
| M | 01/03/2021 | 2.61 | 1.94-3.28 | 0.29 | 0.27-0.32 |

#### **Table S1c. Age-standardised non-covid mortality risks by gender**

| **Gender** | **Date** | **Care or Nursing Home Standardised Risk** | **Care or Nursing Home Confidence Interval** | **Private Home Standardised Risk** | **Private Home Confidence Interval** |
| --- | --- | --- | --- | --- | --- |
| F | 01/02/2019 | 25.96 | 23.26-28.66 | 2.45 | 2.39-2.51 |
| F | 01/03/2019 | 22.35 | 19.82-24.89 | 2.27 | 2.21-2.34 |
| F | 01/04/2019 | 23.34 | 20.92-25.76 | 2.29 | 2.23-2.35 |
| F | 01/05/2019 | 20.27 | 18.07-22.47 | 2.22 | 2.16-2.28 |
| F | 01/06/2019 | 18.92 | 16.75-21.08 | 2.03 | 1.98-2.09 |
| F | 01/07/2019 | 21.13 | 18.79-23.46 | 2.06 | 2-2.12 |
| F | 01/08/2019 | 22.05 | 19.53-24.57 | 2.06 | 2-2.12 |
| F | 01/09/2019 | 21.79 | 19.43-24.14 | 2.05 | 1.99-2.11 |
| F | 01/10/2019 | 26.07 | 23.46-28.68 | 2.24 | 2.18-2.3 |
| F | 01/11/2019 | 27.55 | 24.82-30.27 | 2.32 | 2.26-2.38 |
| F | 01/12/2019 | 28.37 | 25.72-31.03 | 2.68 | 2.62-2.75 |
| F | 01/01/2020 | 27.02 | 24.46-29.58 | 2.69 | 2.62-2.76 |
| F | 01/02/2020 | 24.22 | 21.79-26.64 | 2.28 | 2.22-2.34 |
| F | 01/03/2020 | 27.84 | 25.13-30.55 | 2.48 | 2.42-2.54 |
| F | 01/04/2020 | 35.44 | 32.62-38.26 | 2.41 | 2.35-2.48 |
| F | 01/05/2020 | 21.38 | 19.22-23.55 | 2.04 | 1.98-2.09 |
| F | 01/06/2020 | 17.38 | 15.38-19.39 | 1.97 | 1.91-2.02 |
| F | 01/07/2020 | 18.27 | 16.13-20.41 | 1.99 | 1.93-2.04 |
| F | 01/08/2020 | 19.72 | 17.59-21.86 | 2.03 | 1.97-2.08 |
| F | 01/09/2020 | 19.92 | 17.67-22.17 | 1.99 | 1.94-2.05 |
| F | 01/10/2020 | 22.81 | 20.44-25.18 | 2.15 | 2.09-2.21 |
| F | 01/11/2020 | 20 | 17.86-22.14 | 2 | 1.94-2.06 |
| F | 01/12/2020 | 19.88 | 17.79-21.97 | 2.21 | 2.15-2.27 |
| F | 01/01/2021 | 19.41 | 17.41-21.41 | 2.26 | 2.2-2.32 |
| F | 01/02/2021 | 15.66 | 13.92-17.39 | 1.95 | 1.89-2.01 |
| F | 01/03/2021 | 19.3 | 17.22-21.38 | 2.07 | 2.01-2.12 |
| M | 01/02/2019 | 36 | 32.94-39.06 | 3.33 | 3.24-3.41 |
| M | 01/03/2019 | 31.6 | 28.76-34.44 | 3.2 | 3.11-3.28 |
| M | 01/04/2019 | 31.77 | 28.99-34.55 | 3.27 | 3.19-3.36 |
| M | 01/05/2019 | 28.49 | 25.86-31.11 | 3.06 | 2.98-3.14 |
| M | 01/06/2019 | 32.87 | 29.93-35.81 | 2.84 | 2.76-2.91 |
| M | 01/07/2019 | 27.47 | 24.91-30.04 | 2.94 | 2.86-3.02 |
| M | 01/08/2019 | 29.27 | 26.66-31.87 | 2.85 | 2.77-2.93 |
| M | 01/09/2019 | 31.44 | 28.53-34.34 | 2.88 | 2.8-2.96 |
| M | 01/10/2019 | 32.62 | 29.89-35.34 | 3.16 | 3.07-3.24 |
| M | 01/11/2019 | 37.11 | 34.11-40.11 | 3.35 | 3.27-3.44 |
| M | 01/12/2019 | 41.51 | 38.31-44.71 | 3.71 | 3.62-3.8 |
| M | 01/01/2020 | 40.09 | 37.08-43.11 | 3.68 | 3.59-3.77 |
| M | 01/02/2020 | 33.23 | 30.5-35.96 | 3.21 | 3.13-3.3 |
| M | 01/03/2020 | 38.06 | 35.13-41 | 3.51 | 3.42-3.6 |
| M | 01/04/2020 | 49.74 | 46.42-53.06 | 3.3 | 3.21-3.38 |
| M | 01/05/2020 | 29.72 | 27.03-32.4 | 2.88 | 2.8-2.96 |
| M | 01/06/2020 | 24.28 | 21.82-26.74 | 2.67 | 2.59-2.75 |
| M | 01/07/2020 | 27.45 | 24.81-30.09 | 2.71 | 2.64-2.79 |
| M | 01/08/2020 | 29.58 | 26.87-32.28 | 2.81 | 2.73-2.88 |
| M | 01/09/2020 | 27.38 | 24.88-29.87 | 2.85 | 2.77-2.92 |
| M | 01/10/2020 | 31.53 | 28.9-34.16 | 2.99 | 2.91-3.07 |
| M | 01/11/2020 | 28.34 | 25.73-30.95 | 2.79 | 2.72-2.87 |
| M | 01/12/2020 | 27.98 | 25.36-30.6 | 3.03 | 2.95-3.12 |
| M | 01/01/2021 | 29.44 | 26.88-32 | 3.12 | 3.04-3.21 |
| M | 01/02/2021 | 24.49 | 22.12-26.87 | 2.55 | 2.48-2.62 |
| M | 01/03/2021 | 24.06 | 21.81-26.32 | 2.76 | 2.68-2.84 |

#### **Table S2a. Comparative Mortality Figure (all-cause) comparing Care Homes to Private Homes, by gender**

| **Gender** | **Date** | **Comparative Mortality Risk** | **Confidence Interval** |
| --- | --- | --- | --- |
| F | 01/02/2019 | 10.59 | 9.51-11.81 |
| F | 01/03/2019 | 9.83 | 8.74-11.06 |
| F | 01/04/2019 | 10.2 | 9.16-11.37 |
| F | 01/05/2019 | 9.13 | 8.16-10.22 |
| F | 01/06/2019 | 9.3 | 8.26-10.47 |
| F | 01/07/2019 | 10.26 | 9.15-11.52 |
| F | 01/08/2019 | 10.7 | 9.5-12.04 |
| F | 01/09/2019 | 10.63 | 9.5-11.9 |
| F | 01/10/2019 | 11.63 | 10.47-12.91 |
| F | 01/11/2019 | 11.88 | 10.71-13.18 |
| F | 01/12/2019 | 10.58 | 9.6-11.67 |
| F | 01/01/2020 | 10.05 | 9.1-11.09 |
| F | 01/02/2020 | 10.61 | 9.56-11.78 |
| F | 01/03/2020 | 11.13 | 10.07-12.3 |
| F | 01/04/2020 | 17.52 | 16.38-18.74 |
| F | 01/05/2020 | 13.99 | 12.84-15.24 |
| F | 01/06/2020 | 9.74 | 8.71-10.89 |
| F | 01/07/2020 | 9.76 | 8.68-10.98 |
| F | 01/08/2020 | 9.85 | 8.81-11.01 |
| F | 01/09/2020 | 10.05 | 8.95-11.29 |
| F | 01/10/2020 | 10.72 | 9.67-11.89 |
| F | 01/11/2020 | 10.57 | 9.61-11.63 |
| F | 01/12/2020 | 9.85 | 8.99-10.79 |
| F | 01/01/2021 | 10.3 | 9.53-11.13 |
| F | 01/02/2021 | 8.63 | 7.86-9.48 |
| F | 01/03/2021 | 9.31 | 8.37-10.36 |
| M | 01/02/2019 | 10.82 | 9.89-11.84 |
| M | 01/03/2019 | 9.88 | 8.99-10.87 |
| M | 01/04/2019 | 9.7 | 8.84-10.64 |
| M | 01/05/2019 | 9.31 | 8.45-10.26 |
| M | 01/06/2019 | 11.59 | 10.54-12.75 |
| M | 01/07/2019 | 9.33 | 8.46-10.3 |
| M | 01/08/2019 | 10.26 | 9.34-11.28 |
| M | 01/09/2019 | 10.93 | 9.91-12.05 |
| M | 01/10/2019 | 10.33 | 9.45-11.29 |
| M | 01/11/2019 | 11.06 | 10.15-12.06 |
| M | 01/12/2019 | 11.2 | 10.31-12.16 |
| M | 01/01/2020 | 10.89 | 10.05-11.8 |
| M | 01/02/2020 | 10.34 | 9.47-11.28 |
| M | 01/03/2020 | 10.54 | 9.73-11.42 |
| M | 01/04/2020 | 18.12 | 17.17-19.12 |
| M | 01/05/2020 | 15.24 | 14.2-16.36 |
| M | 01/06/2020 | 9.98 | 9.06-10.98 |
| M | 01/07/2020 | 10.4 | 9.42-11.48 |
| M | 01/08/2020 | 10.67 | 9.69-11.74 |
| M | 01/09/2020 | 9.6 | 8.73-10.56 |
| M | 01/10/2020 | 10.53 | 9.68-11.46 |
| M | 01/11/2020 | 10.63 | 9.81-11.52 |
| M | 01/12/2020 | 10.27 | 9.52-11.08 |
| M | 01/01/2021 | 11.09 | 10.42-11.81 |
| M | 01/02/2021 | 10.13 | 9.33-11 |
| M | 01/03/2021 | 8.74 | 7.97-9.59 |

#### **Table S2b. Comparative Mortality Figure (covid) comparing Care Homes to Private Homes, by gender**

| **Gender** | **Date** | **Comparative Mortality Risk** | **Confidence Interval** |
| --- | --- | --- | --- |
| F | 01/02/2019 |  |  |
| F | 01/03/2019 |  |  |
| F | 01/04/2019 |  |  |
| F | 01/05/2019 |  |  |
| F | 01/06/2019 |  |  |
| F | 01/07/2019 |  |  |
| F | 01/08/2019 |  |  |
| F | 01/09/2019 |  |  |
| F | 01/10/2019 |  |  |
| F | 01/11/2019 |  |  |
| F | 01/12/2019 |  |  |
| F | 01/01/2020 |  |  |
| F | 01/02/2020 |  |  |
| F | 01/03/2020 | 9.48 | 5.59-16.07 |
| F | 01/04/2020 | 24.47 | 21.9-27.34 |
| F | 01/05/2020 | 29.78 | 25.65-34.58 |
| F | 01/06/2020 | 21.56 | 15.72-29.58 |
| F | 01/07/2020 | 38.02 | 23.51-61.5 |
| F | 01/08/2020 | 22.93 | 11.02-47.69 |
| F | 01/09/2020 | 14.11 | 6.69-29.75 |
| F | 01/10/2020 | 12.01 | 8.49-16.99 |
| F | 01/11/2020 | 12.78 | 10.68-15.29 |
| F | 01/12/2020 | 12.73 | 10.78-15.03 |
| F | 01/01/2021 | 12.95 | 11.58-14.49 |
| F | 01/02/2021 | 10.31 | 8.77-12.13 |
| F | 01/03/2021 | 8.98 | 6.35-12.71 |
| M | 01/02/2019 |  |  |
| M | 01/03/2019 |  |  |
| M | 01/04/2019 |  |  |
| M | 01/05/2019 |  |  |
| M | 01/06/2019 |  |  |
| M | 01/07/2019 |  |  |
| M | 01/08/2019 |  |  |
| M | 01/09/2019 |  |  |
| M | 01/10/2019 |  |  |
| M | 01/11/2019 |  |  |
| M | 01/12/2019 |  |  |
| M | 01/01/2020 |  |  |
| M | 01/02/2020 |  |  |
| M | 01/03/2020 | 7.52 | 5.39-10.48 |
| M | 01/04/2020 | 23.31 | 21.52-25.24 |
| M | 01/05/2020 | 36.47 | 32.6-40.8 |
| M | 01/06/2020 | 20.03 | 15.99-25.09 |
| M | 01/07/2020 | 23.06 | 14.52-36.61 |
| M | 01/08/2020 | 25.35 | 12.45-51.63 |
| M | 01/09/2020 | 8.53 | 5.08-14.35 |
| M | 01/10/2020 | 10.44 | 7.97-13.66 |
| M | 01/11/2020 | 11.99 | 10.39-13.84 |
| M | 01/12/2020 | 12.96 | 11.53-14.57 |
| M | 01/01/2021 | 13.29 | 12.19-14.49 |
| M | 01/02/2021 | 11.32 | 9.87-13 |
| M | 01/03/2021 | 8.98 | 6.85-11.77 |

#### **Table S2c. Comparative Mortality Figure (non-covid) comparing Care Homes to Private Homes, by gender**

| **Gender** | **Date** | **Comparative Mortality Risk** | **Confidence Interval** |
| --- | --- | --- | --- |
| F | 01/02/2019 | 10.59 | 9.51-11.81 |
| F | 01/03/2019 | 9.83 | 8.74-11.06 |
| F | 01/04/2019 | 10.2 | 9.16-11.37 |
| F | 01/05/2019 | 9.13 | 8.16-10.22 |
| F | 01/06/2019 | 9.3 | 8.26-10.47 |
| F | 01/07/2019 | 10.26 | 9.15-11.52 |
| F | 01/08/2019 | 10.7 | 9.5-12.04 |
| F | 01/09/2019 | 10.63 | 9.5-11.9 |
| F | 01/10/2019 | 11.63 | 10.47-12.91 |
| F | 01/11/2019 | 11.88 | 10.71-13.18 |
| F | 01/12/2019 | 10.58 | 9.6-11.67 |
| F | 01/01/2020 | 10.05 | 9.1-11.09 |
| F | 01/02/2020 | 10.61 | 9.56-11.78 |
| F | 01/03/2020 | 11.22 | 10.14-12.43 |
| F | 01/04/2020 | 14.68 | 13.49-15.98 |
| F | 01/05/2020 | 10.5 | 9.45-11.68 |
| F | 01/06/2020 | 8.84 | 7.84-9.96 |
| F | 01/07/2020 | 9.19 | 8.14-10.38 |
| F | 01/08/2020 | 9.74 | 8.7-10.9 |
| F | 01/09/2020 | 10 | 8.89-11.24 |
| F | 01/10/2020 | 10.61 | 9.52-11.82 |
| F | 01/11/2020 | 10 | 8.94-11.18 |
| F | 01/12/2020 | 8.99 | 8.06-10.03 |
| F | 01/01/2021 | 8.58 | 7.71-9.55 |
| F | 01/02/2021 | 8.03 | 7.16-9.01 |
| F | 01/03/2021 | 9.34 | 8.35-10.45 |
| M | 01/02/2019 | 10.82 | 9.89-11.84 |
| M | 01/03/2019 | 9.88 | 8.99-10.87 |
| M | 01/04/2019 | 9.7 | 8.84-10.64 |
| M | 01/05/2019 | 9.31 | 8.45-10.26 |
| M | 01/06/2019 | 11.59 | 10.54-12.75 |
| M | 01/07/2019 | 9.33 | 8.46-10.3 |
| M | 01/08/2019 | 10.26 | 9.34-11.28 |
| M | 01/09/2019 | 10.93 | 9.91-12.05 |
| M | 01/10/2019 | 10.33 | 9.45-11.29 |
| M | 01/11/2019 | 11.06 | 10.15-12.06 |
| M | 01/12/2019 | 11.2 | 10.31-12.16 |
| M | 01/01/2020 | 10.89 | 10.05-11.8 |
| M | 01/02/2020 | 10.34 | 9.47-11.28 |
| M | 01/03/2020 | 10.84 | 9.98-11.77 |
| M | 01/04/2020 | 15.08 | 14.02-16.22 |
| M | 01/05/2020 | 10.32 | 9.38-11.36 |
| M | 01/06/2020 | 9.1 | 8.18-10.12 |
| M | 01/07/2020 | 10.12 | 9.15-11.2 |
| M | 01/08/2020 | 10.54 | 9.57-11.61 |
| M | 01/09/2020 | 9.62 | 8.74-10.59 |
| M | 01/10/2020 | 10.55 | 9.65-11.52 |
| M | 01/11/2020 | 10.14 | 9.2-11.18 |
| M | 01/12/2020 | 9.22 | 8.36-10.17 |
| M | 01/01/2021 | 9.42 | 8.59-10.33 |
| M | 01/02/2021 | 9.61 | 8.67-10.64 |
| M | 01/03/2021 | 8.72 | 7.9-9.62 |

#### **Table S3. Demographic and Clinical Characteristics of Care Home and Private Home Residents on the 1st of February 2020**

|  |  | **Overall** | | **Care or Nursing Home** | | **Private Home** | |
| --- | --- | --- | --- | --- | --- | --- | --- |
|  |  | **N** | **%** | **N** | **%** | **N** | **%** |
| **Total** |  | 4410388 | 100 | 101055 | 100 | 4309333 | 100 |
| Care Home Type | Care Home | 52630 | 1.19 | 52630 | 52.08 |  |  |
|  | Care or Nursing Home | 2422 | 0.05 | 2422 | 2.4 |  |  |
|  | Nursing Home | 46003 | 1.04 | 46003 | 45.52 |  |  |
|  | Private Home | 4309333 | 97.71 |  |  | 4309333 | 100 |
| Gender | Female | 2372622 | 53.8 | 70634 | 69.9 | 2301988 | 53.42 |
|  | Male | 2037766 | 46.2 | 30421 | 30.1 | 2007345 | 46.58 |
| Age in Years | Mean, SD | 75 | 7.52 | 86 | 7.88 | 75 | 7.32 |
| Self - reported Ethnicity | Asian or British Asian | 118874 | 2.7 | 600 | 0.59 | 118274 | 2.74 |
|  | Black | 34907 | 0.79 | 483 | 0.48 | 34424 | 0.8 |
|  | Missing | 1019287 | 23.11 | 24610 | 24.35 | 994677 | 23.08 |
|  | Mixed | 12818 | 0.29 | 202 | 0.2 | 12616 | 0.29 |
|  | Other | 28386 | 0.64 | 351 | 0.35 | 28035 | 0.65 |
|  | White | 3196116 | 72.47 | 74809 | 74.03 | 3121307 | 72.43 |
| Geographical Region | East | 1039428 | 23.57 | 23393 | 23.15 | 1016035 | 23.58 |
|  | East Midlands | 781700 | 17.72 | 18514 | 18.32 | 763186 | 17.71 |
|  | London | 169793 | 3.85 | 1843 | 1.82 | 167950 | 3.9 |
|  | North East | 202664 | 4.6 | 4476 | 4.43 | 198188 | 4.6 |
|  | North West | 409382 | 9.28 | 9913 | 9.81 | 399469 | 9.27 |
|  | South East | 325023 | 7.37 | 8362 | 8.27 | 316661 | 7.35 |
|  | South West | 740911 | 16.8 | 16992 | 16.81 | 723919 | 16.8 |
|  | West Midlands | 155270 | 3.52 | 3109 | 3.08 | 152161 | 3.53 |
|  | Yorkshire and The Humber | 585269 | 13.27 | 14441 | 14.29 | 570828 | 13.25 |
|  | Missing | 948 | 0.02 | 12 | 0.01 | 936 | 0.02 |
| Quintile of Index of Multiple Deprivation | 1 - Least Deprived | 623477 | 14.14 | 18408 | 18.22 | 605069 | 14.04 |
|  | 2 | 772496 | 17.52 | 20635 | 20.42 | 751861 | 17.45 |
|  | 3 | 991751 | 22.49 | 22333 | 22.1 | 969418 | 22.5 |
|  | 4 | 1010024 | 22.9 | 20470 | 20.26 | 989554 | 22.96 |
|  | 5 - Most Deprived | 985648 | 22.35 | 18589 | 18.39 | 967059 | 22.44 |
|  | Missing | 26992 | 0.61 | 620 | 0.61 | 26372 | 0.61 |
| History of Stroke |  | 295451 | 6.7 | 21898 | 21.67 | 273553 | 6.35 |
| Dementia |  | 214471 | 4.86 | 60021 | 59.39 | 154450 | 3.58 |
| Diabetes |  | 985697 | 22.35 | 25044 | 24.78 | 960653 | 22.29 |
| Chronic Kidney Disease |  | 449459 | 10.19 | 20971 | 20.75 | 428488 | 9.94 |
| Cancer |  | 665420 | 15.09 | 16640 | 16.47 | 648780 | 15.06 |
| Chronic Liver Disease |  | 39473 | 0.9 | 1023 | 1.01 | 38450 | 0.89 |
| Chronic Cardiac Disease |  | 880098 | 19.96 | 31031 | 30.71 | 849067 | 19.7 |
| Chronic Respiratory Disease |  | 500848 | 11.36 | 13588 | 13.45 | 487260 | 11.31 |

#### **Table S4. Demographic and Clinical Characteristics of Care Home and Private Home Residents on the 1st of February 2021**

|  |  | **Overall** | | **Care or Nursing Home** | | **Private Home** | |
| --- | --- | --- | --- | --- | --- | --- | --- |
|  |  | **N** | **%** | **N** | **%** | **N** | **%** |
| **Total** |  | 4452375 | 100 | 95295 | 100 | 4357080 | 100 |
| Care Home Type | Care Home | 50183 | 1.13 | 50183 | 52.66 |  |  |
|  | Care or Nursing Home | 2128 | 0.05 | 2128 | 2.23 |  |  |
|  | Nursing Home | 42984 | 0.97 | 42984 | 45.11 |  |  |
|  | Private Home | 4357080 | 97.86 |  |  | 4357080 | 100 |
| Gender | Female | 2393928 | 53.77 | 67592 | 70.93 | 2326336 | 53.39 |
|  | Male | 2058447 | 46.23 | 27703 | 29.07 | 2030744 | 46.61 |
| Age in Years | Mean, SD | 75 | 7.47 | 86 | 8.01 | 75 | 7.29 |
| Self - reported Ethnicity | Asian or British Asian | 123749 | 2.78 | 627 | 0.66 | 123122 | 2.83 |
|  | Black | 36002 | 0.81 | 451 | 0.47 | 35551 | 0.82 |
|  | Missing | 1012050 | 22.73 | 21728 | 22.8 | 990322 | 22.73 |
|  | Mixed | 13455 | 0.3 | 200 | 0.21 | 13255 | 0.3 |
|  | Other | 29913 | 0.67 | 320 | 0.34 | 29593 | 0.68 |
|  | White | 3237206 | 72.71 | 71969 | 75.52 | 3165237 | 72.65 |
| Geographical Region | East | 1049954 | 23.58 | 21922 | 23 | 1028032 | 23.59 |
|  | East Midlands | 787862 | 17.7 | 17415 | 18.27 | 770447 | 17.68 |
|  | London | 172211 | 3.87 | 1739 | 1.82 | 170472 | 3.91 |
|  | North East | 203248 | 4.56 | 4153 | 4.36 | 199095 | 4.57 |
|  | North West | 413879 | 9.3 | 9553 | 10.02 | 404326 | 9.28 |
|  | South East | 328073 | 7.37 | 7716 | 8.1 | 320357 | 7.35 |
|  | South West | 751365 | 16.88 | 16466 | 17.28 | 734899 | 16.87 |
|  | West Midlands | 154524 | 3.47 | 2928 | 3.07 | 151596 | 3.48 |
|  | Yorkshire and The Humber | 590256 | 13.26 | 13378 | 14.04 | 576878 | 13.24 |
|  | Missing | 1003 | 0.02 | 25 | 0.03 | 978 | 0.02 |
| Quintile of Index of Multiple Deprivation | 1 - Least Deprived | 622464 | 13.98 | 17312 | 18.17 | 605152 | 13.89 |
|  | 2 | 776306 | 17.44 | 19592 | 20.56 | 756714 | 17.37 |
|  | 3 | 1000070 | 22.46 | 21033 | 22.07 | 979037 | 22.47 |
|  | 4 | 1021600 | 22.95 | 19681 | 20.65 | 1001919 | 23 |
|  | 5 - Most Deprived | 998254 | 22.42 | 17003 | 17.84 | 981251 | 22.52 |
|  | Missing | 33681 | 0.76 | 674 | 0.71 | 33007 | 0.76 |
| History of Stroke |  | 292461 | 6.57 | 20182 | 21.18 | 272279 | 6.25 |
| Dementia |  | 194738 | 4.37 | 54435 | 57.12 | 140303 | 3.22 |
| Diabetes |  | 1017418 | 22.85 | 24431 | 25.64 | 992987 | 22.79 |
| Chronic Kidney Disease |  | 402903 | 9.05 | 17392 | 18.25 | 385511 | 8.85 |
| Cancer |  | 673175 | 15.12 | 15398 | 16.16 | 657777 | 15.1 |
| Chronic Liver Disease |  | 41425 | 0.93 | 1073 | 1.13 | 40352 | 0.93 |
| Chronic Cardiac Disease |  | 871017 | 19.56 | 28623 | 30.04 | 842394 | 19.33 |
| Chronic Respiratory Disease |  | 493455 | 11.08 | 12707 | 13.33 | 480748 | 11.03 |
